# Phenotypic markers misclassify isolates of *Salmonella typhimurium* as *S. typhi*, but were correctly identified by whole genome sequencing during a septicemia study in western Kenya

**DOI:** 10.1101/2025.02.28.25323083

**Authors:** Joseph Kaingu, Kimita Gathii, Carolyne Kifude, James Nonoh, Lillian Ogonda, Amos Onditi, Kirti Tiwari, John Waitumbi

## Abstract

Antimicrobial resistance (AMR) is a rising global health threat and estimated to cause 700,000 deaths annually. Although blood cultures (BCs) are the reference standard to diagnose bloodstream infections and inference of antimicrobial susceptibility testing (AST), the method could fail to differentiate bacteria with similar biochemical characteristics. This study evaluated the utility of whole genome sequencing (WGS) in complimenting BCs in bacteria identification and assessing AMR. Blood samples came from septicemic patients attending county referral hospitals in western Kenya and around the Lake Victoria region. BCs and AST were performed on BD Bactec 9050 and Phoenix 100 respectively. Out of the 960 BCs, 17 had uncontaminated growth and were evaluated by WGS on the Oxford Nanopore PromethION platform. BD Phoenix system identified the 17 bacteria isolates as: 4 *Escherichia coli*, 8 *Salmonella enterica* serovar Typhi, 1 unspeciated *Salmonella*, 3 *Staphylococcus aureus* and 1 *Streptococcus pneumoniae*. WGS results differed from BCs in identifying *Salmonella species*, with WGS identifying the species as *Salmonella enterica* serovar Typhimurium. Conversely, WGS detected AMR genes in bacteria that AST had classified as susceptible. In conclusion, we caution that BCs may not be providing correct identity of *Salmonella* species. The observed discrepancies between phenotypic and genotypic markers of drug resistance highlight the challenges in interpreting and predicting the functional utility of AMR genes.

## INTRODUCTION

Blood cultures (BCs) serve as the gold standard for diagnosing bloodstream infections and when coupled with antimicrobial susceptibility testing (AST), provide the associated antimicrobial resistance (AMR) profile. AMR is a major global public health challenge which threatens to render antibiotic management strategies ineffective [1]. Sub-Saharan Africa shoulder’s the highest morbidity and mortality burden, and the associated high AMR treatment costs [2, 3]. The misuse of antibiotics in humans and animals is the major driver of AMR, and leads to poor infection control, environmental contamination, and geographical spread of resistant bacteria [4]. The identification of AMR genes can supplement traditional phenotypic antimicrobial susceptibility testing (AST) by predicting bacterial resistance to specific antibiotics, helping to guide the selection of effective treatments [5].

Correct identification of bacteria species is important for proper attribution and for epidemiological tracking. Unfortunately, certain bacteria exhibit similar biochemical characteristics, making differentiation through traditional culture methods challenging. In such cases, molecular techniques like PCR-based identification and sequencing of the 16S rRNA gene are employed for accurate identification [5, 6]. For example, identifying *Salmonella typhimurium* from other *Salmonella enterica* serovars using phenotypic or chemical properties can be challenging due to their similarities in biochemical characteristics [7]. Although serotyping is highly effective in distinguishing between the two sub-species of *Salmonella*, it is not routinely done [8]. In addition, some commercial systems do not have *S. typhimurium* or misdiagnose it, leading to under-recognition [7].

There are several genotypic methods for the detection of antimicrobial resistance (AMR) genes in bacteria. These genotypic methods are essential tools in microbiology, helping to inform treatment decisions, track resistance trends, and understand the genetic mechanisms underlying antimicrobial resistance. The choice of method often depends on the specific context, including the required sensitivity, specificity, speed, and resources available. Multiplex PCR assays that amplify specific AMR genes have high sensitivity and specific and can detect multiple genes simultaneously [9]. In Kenya, most hospitals use GeneXpert [10] for mycobacterium AMR gene detection [11]. But the test has broader capability, including detection of carbapenemases [10] and vanA/vanB [12]. Of the molecular methods, next generation whole genome sequencing (WGS) is the most robust, has high-throughput capability that allows for the simultaneous detection of multiple genes or entire genomes, thus can provide comprehensive data on AMR genes, including novel mutations and plasmid content [13, 14]. Using bioinformatics tools and reference databases, WGS is able to accurately identify bacteria, determine the AMR genes they carry, including resistance mechanisms [15, 16]. This study evaluated the utility of WGS in calling bacteria species and AMR genes in blood cultures obtained from septicemic patients that had prior AST results.

## MATERIALS AND METHODS

### Study site and ethical approval

This study was part of an antimicrobial resistance surveillance study in septicemic patients that was conducted at the Walter Reed Army Institute of Research-Africa (WRAIR-A), Kombewa Center for Clinical Research (CCR), Kisumu Field Station. The samples were collected between March 2022 and April 2023 from county referral hospitals in western Kenya and around the Lake Victoria region (Busia, Kakamega, Homabay, Kisumu and Vihiga). The study was approved by the Kenya Medical Research Institute Scientific and Ethics Review Unit (KEMRI SERU, # 4246) and the WRAIR Human Subject Protection Branch, (WRAIR #2888). Approval for waiver of consent was obtained as blood cultures are deemed routine and the microbiology results from the study are used for clinical management of individual patients-SERU ethical approval on August 10, 2021.

### Culture and phenotypic antimicrobial susceptibility testing

960 blood cultures (BCs) were collected from patients suspected of having septicemia and incubated in BD Bactec 9050 (Sparks, Maryland, USA). BCs with growth were sub-cultured on blood agar, chocolate blood agar, and MacConkey agar. Following Gram staining, the bacteria identification and AST profiles were performed using the BD Phoenix 100 system with software v6.01 and Phoenix Update Disk (PUD) v5.11, according to the World Health Organization - Global antimicrobial resistance and use surveillance (GLASS) report [17].

### Whole genome sequencing

Bacterial cultures for sequencing were subcultured for purity in 5% Columbia blood agar (Thermo Fisher Diagnostics) and incubated overnight at 37°C. A colony was picked with a loop and added to 250 μL of phosphate buffered saline. Genomic DNA was extracted using the ZymoBIOMICS DNA/RNA Miniprep Kit (Zymo RESEARCH, California, US), according to the manufacturer’s instructions. Genomic DNA library was prepared for sequencing using the Native Barcoding Kit 24 v14 (SQK-NBD114.24) (Oxford Nanopore Technologies, Oxford, UK), according to the manufacturer’s instructions. 200 μL of the prepared DNA library was loaded into an R10.4.1 flow cell and then sequenced on the PromethION 2 Solo instrument (Oxford Nanopore Technologies, Oxford, UK) for a maximum of 72 hours. Raw pod5 files from the PromethION were base called using Dorado v0.5.2 (https://github.com/nanoporetech/dorado). The demultiplexed sequences were then run on EDGE Bioinformatics v2.4.0 Pipeline [18], that incorporates quality control using FaQCs v1.34 [19]; assembly and annotation using Flye v.2.9.3 [20]; and taxonomic classification using Kraken v1.2.0 [21] to identify the bacteria. The Flye assembled contigs were further analyzed for AMR genes using abritAMR v1.0.13 [22], which uses the National Center for Biotechnology Information (NCBI)’s AMRFinderPlus database [23] and the online version of the Resistance Gene Identifier (RGI) v6.0.3 (https://card.mcmaster.ca/analyze/rgi) from the Comprehensive Antimicrobial Resistance Database (CARD) v3.2.8 [24]. The abritAMR’s ≥ 90% reference sequence identity and coverage threshold were used in both tools (Sherry et al., 2023). An isolate was defined as genotypic resistant by the presence of at least one AMR gene or chromosomal point mutation (PM) known to confer resistance to a given antimicrobial agent and as genotypic susceptible when no ARG or PM was found [25, 26].

### Confirmation of the identity of *Salmonella* isolates by phylogenetic analysis

To confirm the identity of our study *Salmonella* isolates, phylogenetic analysis was done with both *S. typhimurium* and *S. typhi* global sequences downloaded from the NCBI database (https://blast.ncbi.nlm.nih.gov) and Enterobase database v1.2.0 (https://enterobase.warwick.ac.uk) [27]. The Parsnp v2.0.3 [28] was used for phylogenetic analysis. The created tree was visualized in iTOL v6.9 [29].

## RESULTS

### Blood cultures failed to identify *Salmonella typhimurium*

Out of the 960 blood cultures, 123 (12.8%) had bacterial growth, and of these, only 17 (13.8%) were usable (not contaminated). Phenotypically, the bacteria were classified as 4 Gram positives: *S. aureus* (n=3), and *S. pneumoniae* (n=1) and 13 Gram negatives: *E. coli* (n=4), *S. typhi*, (n=8), and unspeciated *Salmonella* (n=1). WGS was concordant with phenotypic identification of all other bacteria, apart from *Salmonella spp*. By WGS, the *Salmonella* isolates were identified as *Salmonella enterica* serovar *typhimurium*. By phylogenetic analysis, the *Salmonella* study genomes formed a single cluster with the global *Salmonella enterica* serovar *typhimurium* sequences (**Figure 1**, red fonts).

**Figure 1.**
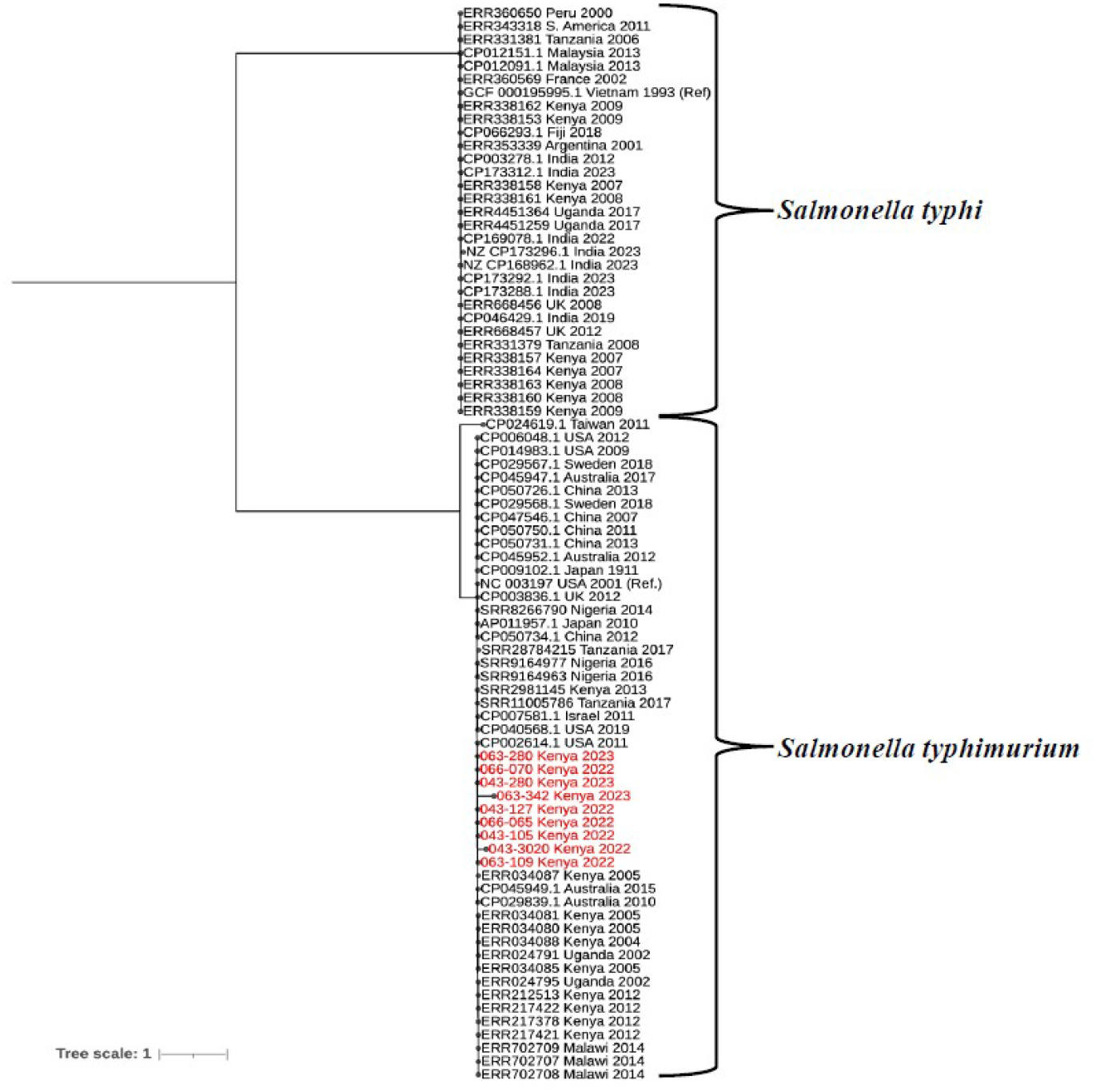
Phylogenetic tree generated from *Salmonella* core genome single nucleotide polymorphisms using Parsnp v2.0.3. The study genomes (highlighted in red) clustered with *S. typhimurium* and not *S. typhi*. GCF_0001959951=*S. typhi* reference genome; NC_003197=*S. typhimurium* reference genome

### Antimicrobial resistance profiles

Detailed phenotypic resistance profiles for *Salmonella spp*., *E. coli, S. aureus* and *S. pneumonia* are shown in **Table 1**

**Table 1:**
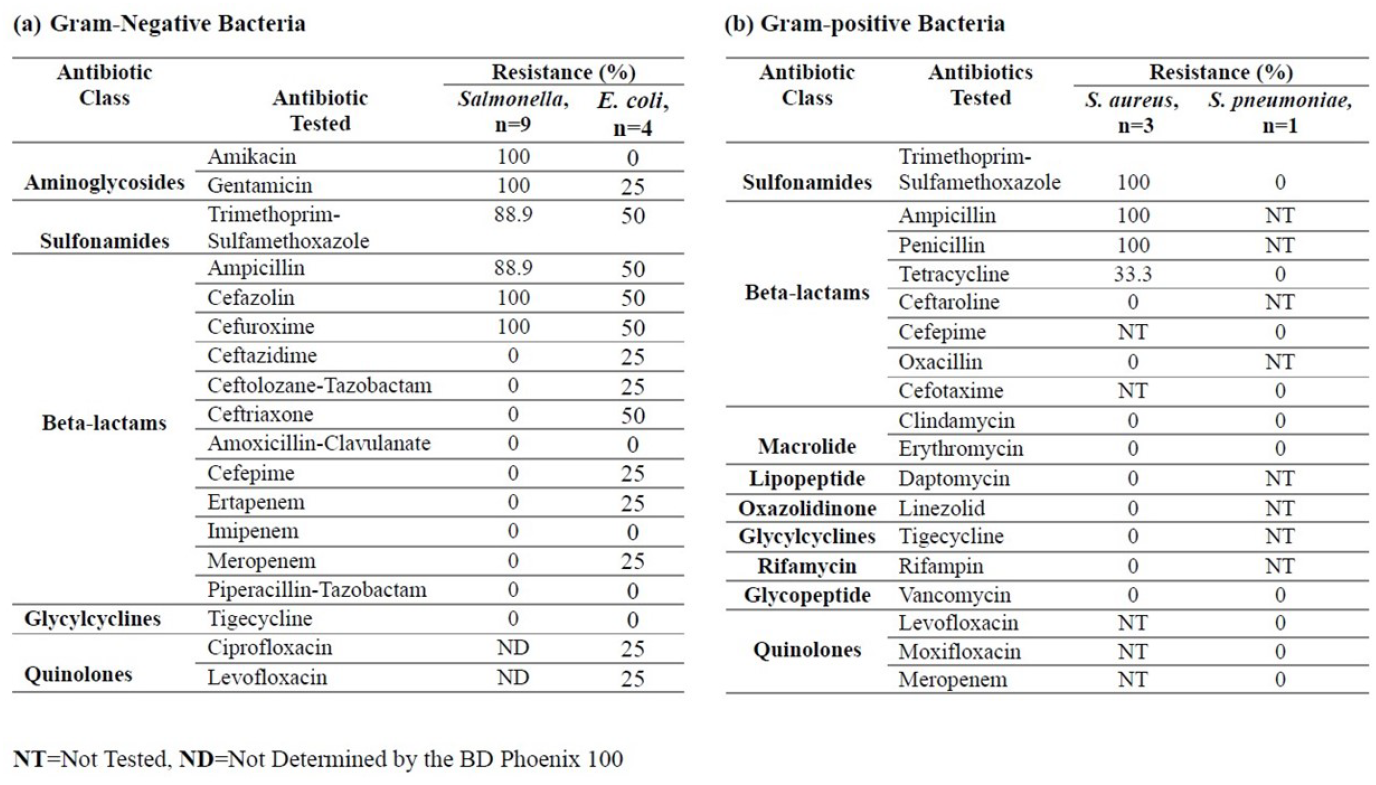
Antibiotic resistance profile for *Salmonella spp*., *Escherichia coli, Staphylococcus aureus* and *Streptococcus pneumoniae* were performed in the BD Phoenix 100 system.

Figure 2 shows the phenotypic and genotypic AMR for the study isolates. All the nine isolates of *Salmonella* (**Panel A**) showed 100% resistance to aminoglycosides (amikacin and gentamycin) and sulfonamide (trimethoprim-sulfamethoxazole). The isolates had 100% sensitivity to betalactams, except ampicillin, cefazolin, and cefuroxime that had 88.9%, 100% and 100% resistance respectively and 100% sensitivity to glycylcycline (tigecycline). There was a higher phenotype and genotype concordance in the resistance profile to aminoglycosides. AMR genes that code for beta-lactams resistance were identified in both the susceptible and resistant isolates. Despite the *Salmonella* isolates being susceptible to tigecycline, multiple AMR genes were identified. For sulfonamides, only the ***rsm****A* resitance gene was present in all the *Salmonella* isolates.

In all instances except one, the *E. coli* isolates (**Figure 2, Panel B**) were 100% sensitive aminoglycosides, but had variable resistance to beta-lactams, sulfonamide and quinolones and 100% sensitivity to glycylcycline (tigecycline). Although *E. coli* isolates showed high sensitivity to aminoglycosides, multiple AMR genes that code for resistance to amonoglycosides were identified. Likewise, AMR genes that code for beta-lactams resistance were identified in both the susceptible and resistant *E. coli*. Similar to the *Salmonella* isolates, multiple AMR genes that code for resistance to tigecycline were identified despite the susceptible phonotype. For sulfonamides, only the *rsmA* resistance gene was present in all the *E. coli* isolates. AMR genes that confer resistance to quinolones were present in nearly all the *E. coli* isolates.

**Figure 2:**
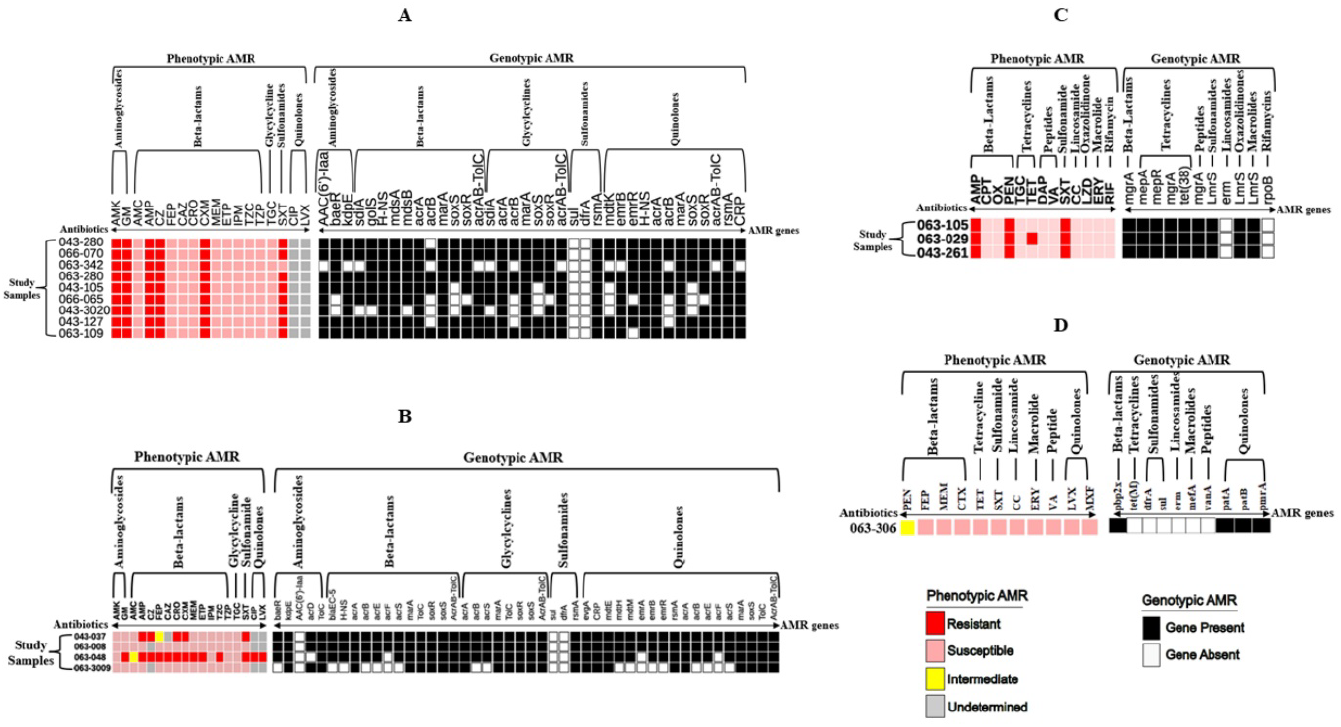
Phenotypic and genotypic AMR profiles of the study isolates: *Salmonella spp*. (Panel A), *Escherichia coli E. coli* (Panel B), *Staphyloccucus aureus* (Panel C), and *Streptococcus pneumoniae* (Panel D) to the following antibiotics, Aminogylcosides (Amikacin= AMK, Gentamicin=GM); Beat-lactams (Amoxicillin-Clavulanate=AMC, Ampicillin=AMP, Cefazolin=CZ, Ceftazidime=CAZ, Ceftriaxone=CRO, Cefuroxime=CXM, Meropenem=MEM, Ertapenem=ETP, Imipenem=IPM, Ceftolozane-Tazobactam=TZC, Ceftaroline=CPT, Oxacillin=OX, TZP=Piperacillin-Tazobactam); Glycylcycline (Tigecycline=TGC); Sulfonamides (Trimethoprim-Sulfamethoxazole=SXT) Quinolones (Ciprofloxacin=CIP, Levofloxacin=LVX).

By culture and sensitivity, the *S. aureus* isolates **(Figure 2, Panel C)** were completely resistant to two beta-lactams (ampicillin and penicillin) and sulfonamide. One isolate was resistant to tetracycline. The isolates were completely susceptible to the other antibiotics tested (ceftaroline, oxacillin tigecycline, daptomycin, vancomycin, linezolid, clindamycin, erythromycin and rifamycin). The AMR presence/absence of genes did not match the expected phenotype profile. Unlike the *Salmonella* isolates, it was the *LmrS* gene that code for sulfonamide resistance that was identified in all the *S. aureus* isolates, and not *rsmA* gene.

The phenotypic and genotypic profile of the lone isolate of *S. pneumoniae* is shown in **Figure 2**, (**Panel D**). The isolate was susceptible to all the tested antibiotics. AMR genes coding for resistance to beta-lactams and quinolones were identified, despite the isolates being sensitive to these antibiotics. In concordance to AST profile, AMR genes coding for resistance to tetrcyclines, sulfonamides, lincosamide, macrolide and peptides were absent.

## DISCUSSION

Bacterial infections in the bloodstream are a major cause of sepsis and fatalities, making precise identification of the causative agents and their antimicrobial resistance (AMR) profiles essential for effective management [3, 30].

In this study, phenotypic and genotypic markers by AST and WGS respectively were concordant in identification of the bacterial isolates, except for the assignment of *Salmonella* serovars. Phenotypic markers identified them as either *Salmonella spp*. or *S. typhi*, while genotypic markers identified them as *S. typhimurium. Salmonella enterica* serovars have similar biochemical characteristic that make it difficult to distinguish them [7]. We think the genotype classification is correct for several reasons. One, WGS has high discriminative power, and in previous studies it had been found to be more accurate in the identification of bacteria compared to four automated microbiology systems, including the BD Phoenix 100 that was used in the current study [31]. Two, as shown in **Figure 1**, the 9 *Salmonella* study genomes clustered with the global *S. typhimurium*, and not *S. typhi*. Three, we noted that the BD Phoenix 100 bacterial ID list [32] has *S. typhi* but no *S. typhimurium*. Misclassification of *S. typhimurium* as *S. typhi* can lead to underestimation of the burden of *S. typhimurium* and also because *Salmonella enterica* serovars have distinct clinical presentations, epidemiology, and treatment requirements [33]. In humans, *S. typhimurium* typically causes gastroenteritis, characterized by diarrhea, abdominal pain, fever, and vomiting. In severe cases, it can lead to systemic infection, particularly in immunocompromised individuals. [34, 35]. In contrast, *S. typhi* causes a systemic infection affecting multiple organs [36].

As illustrated in **Figure 2 (Panel A)**, *S. typhimurium* showed 100% sensitivity to beta-lactams, apart from ampicillin, cefazolin, and cefuroxime. Previous studies in Kenya had showed non-sensitivity to most beta-lactams [37–39]. AMR genes that code for beta-lactams resistance were identified in both the susceptible and resistant isolates. Unlike the beta-lactams, there was a better phenotype and genotype concordance in the resistance profile to aminoglycosides (amikacin and gentamicin). Similar to our study, several studies have found trimethoprim-sulfamethoxazole resistance in *S. typhimurium* which was attributed to *sul* and *dfrA* AMR genes [39–41]. In our study, we only identified the *rsmA* gene that is known to confer resistance to trimethoprim-sulfamethoxazole. Similarly, AMR genes that code for glycylicyclines (tigecycline) were identified despite all *S. typhimurium* isolates being phenotypically susceptible.

*E. coli*, which mostly resides in the gut of humans and animals, is mainly transmitted through consumption of contaminated food and water, and has the ability to survive and reproduce in the environment [42]. As shown in **Figure 2 (Panel B)**, *E. coli* isolates were phenotypically sensitive to aminoglycosides (amikacin and gentamicin), suggesting their potential as effective treatment options. The observed phenotypic resistance to beta-lactams, sulfonamides (e.g., trimethoprim-sulfamethoxazole), and quinolones aligns with the widespread prevalence of AMR genes for these drug classes. Similar to *S. typhimurium* observed presence of the *rsmA* resistance gene across all gram negative isolates may explain the observed resistance to sulfonamides. This gene likely plays a key role in resistance to this class of antibiotics. The phenotypic susceptibility to tigecycline is promising, suggesting its potential as a robust treatment option for *E. coli* infections. However, the detection of multiple AMR genes associated with tigecycline resistance is concerning.

The phenotypic and genotype profiles of the three isolates of *S. aureus* are shown in **Figure 4 (Panel C)**. The isolates were completely resistant to ampicillin and penicillin, but susceptible to ceftaroline and oxacillin. The isolates were also resistant to trimethoprim-sulfamethoxazole and had variable resistance to tetracycline. The phenotypic resistance to beta-lactams (ampicillin and penicillin), sulfonamides (trimethoprim-sulfamethoxazole) and tetracycline is similar to previous studies done in Kenya [43–46]. Irrespective of phenotypic resistance profiles, AMR genes were identified in all isolates apart from lincosamide and rifamycin.

*S. pneumoniae* was susceptible to all the tested antibiotics (**Figure 2, Panel D**). Resistance genes were only found for beta-lactams, and quinolones, and none for tetracyclines, sulfonamides, lacosamides, macrolides and peptides. In general, the phenotype profile agrees with a systematic review by Droz et al that found high susceptibility of *S. pneumoniae* to beta-lactams in Africa [47]. In another study done in Kenya [48], *S. pneumoniae* was found to have increased resistance to oxacillin and erythromycin, which is contrary to our findings. Lastly, our study had only one isolate of *S. pneumoniae* and hence making it difficult to generalize these findings.

In conclusion, phenotypic markers failed to correctly identify the serovars of *Salmonella enterica* and classified them as *Salmonella spp*. or *S. typhi*. WGS correctly identified the misclassified *Salmonella* as *S. t*yphimurium. Additionally, our results highlight the importance of integrating genotypic and phenotypic data for a comprehensive understanding of AMR. Phenotypic sensitivity testing alone may underestimate the potential for resistance development, especially when silent or partially expressed AMR genes are present. Continued surveillance of AMR genes, even in phenotypically susceptible isolates, is critical to predict and mitigate emerging resistance threats. The results underscore the need for combination therapies or alternative treatment approaches to address variable resistance profiles, especially for beta-lactams, sulfonamides, and quinolones.

## Data Availability

The datasets used and/or analysed during the current study are available from the corresponding author on reasonable request.

## Disclaimer

Material has been reviewed by the Walter Reed Army Institute of Research. There is no objection to its publication. The opinions or assertions contained herein are the private views of the author, and they are not to be construed as official, or as reflecting true views of the Department of the Army or the Department of Defense. The investigators have adhered to the policies for protection of human subjects as prescribed in AR 70–25.

## REFERENCES

[1] Aljeldah MM. Antimicrobial Resistance and Its Spread Is a Global Threat. Epub ahead of print 2022. DOI: 10.3390/antibiotics11081082.

[2] Godman B, Egwuenu A, Wesangula E, et al. Tackling antimicrobial resistance across sub-Saharan Africa: current challenges and implications for the future. Expert Opin Drug Saf 2022; 21: 1089–1111.

[3] Seni J, Mwakyoma AA, Mashuda F, et al. Deciphering risk factors for blood stream infections, bacteria species and antimicrobial resistance profiles among children under five years of age in North-Western Tanzania: A multicentre study in a cascade of referral health care system. BMC Pediatr; 19. Epub ahead of print 26 January 2019. DOI: 10.1186/s12887-019-1411-0.

[4] McEwen SA, Collignon PJ. Antimicrobial Resistance: a One Health Perspective. Microbiol Spectr; 6. Epub ahead of print 6 April 2018. DOI: 10.1128/microbiolspec.arba-0009-2017.

[5] Yamin D, Uskoković V, Wakil AM, et al. Current and Future Technologies for the Detection of Antibiotic-Resistant Bacteria. Diagnostics 2023; 13: 1–43.

[6] Galluzzi L, Magnani M, Saunders N, et al. Current molecular techniques for the detection of microbial pathogens. Sci Prog 2007; 90: 29–50.

[7] Awang MS, Bustami Y, Hamzah HH, et al. Advancement in salmonella detection methods: From conventional to electrochemical-based sensing detection. Biosensors 2021; 11: 1–42.

[8] Diep B, Barretto C, Portmann A, et al. Salmonella Serotyping ; Comparison of the Traditional Method to a Microarray-Based Method and an in silico Platform Using Whole Genome Sequencing Data. 10. Epub ahead of print 2019. DOI: 10.3389/fmicb.2019.02554.

[9] Anjum MF, Zankari E, Hasman H. Molecular Methods for Detection of Antimicrobial Resistance. Microbiol Spectr; 5. Epub ahead of print 2017. DOI: 10.1128/microbiolspec.arba-0011-2017.

[10] Ding L, Shi Q, Han R, et al. Comparison of Four Carbapenemase Detection Methods for bla KPC-2 Variants . Microbiol Spectr 2021; 9: 1–10.

[11] Bwana P, Ageng’o J, Mwau M. Performance and usability of Cepheid GeneXpert HIV-1 qualitative and quantitative assay in Kenya. PLoS One 2019; 14: 1–10.

[12] Li ZL, Luo QB, Xiao SS, et al. Evaluation of genexpert vana/vanb in the early diagnosis of vancomycin-resistant enterococci infection. PLoS Negl Trop Dis 2021; 15: 1–12.

[13] Tamma PD, Fan Y, Bergman Y, et al. Applying rapid whole-genome sequencing to predict phenotypic antimicrobial susceptibility testing results among carbapenem-resistant klebsiella pneumoniae clinical isolates. Antimicrob Agents Chemother 2019; 63: 1–12.

[14] Salam MA, Al-Amin MY, Pawar JS, et al. Conventional methods and future trends in antimicrobial susceptibility testing. Saudi J Biol Sci 2023; 30: 103582.

[15] Sydenham T V., Overballe-Petersen S, Hasman H, et al. Complete hybrid genome assembly of clinical multidrug-resistant bacteroides fragilis isolates enables comprehensive identification of antimicrobial-resistance genes and plasmids. Microb Genomics 2019; 5: 1–18.

[16] Su M, Satola SW, Read TD. Genome-based prediction of bacterial antibiotic resistance. J Clin Microbiol; 57. Epub ahead of print 2019. DOI: 10.1128/JCM.01405-18.

[17] GLASS. Global Antimicrobial Resistance and Use Surveillance System (GLASS) Report 2021. 2021.

[18] Li PE, Lo CC, Anderson JJ, et al. Enabling the democratization of the genomics revolution with a fully integrated web-based bioinformatics platform. Nucleic Acids Res 2017; 45: 67– 80.

[19] Lo CC, Chain PSG. Rapid evaluation and quality control of next generation sequencing data with FaQCs. BMC Bioinformatics 2014; 15: 1–8.

[20] Kolmogorov M, Yuan J, Lin Y, et al. Assembly of long, error-prone reads using repeat graphs. Nat Biotechnol 2019; 37: 540–546.

[21] Wood DE, Salzberg SL. Kraken: Ultrafast metagenomic sequence classification using exact alignments. Genome Biol; 15. Epub ahead of print 2014. DOI: 10.1186/gb-2014-15-3-r46.

[22] Sherry NL, Horan KA, Ballard SA, et al. An ISO-certified genomics workflow for identification and surveillance of antimicrobial resistance. Nat Commun; 14. Epub ahead of print 2023. DOI: 10.1038/s41467-022-35713-4.

[23] Feldgarden M, Brover V, Fedorov B, et al. Curation of the AMRFinderPlus databases: Applications, functionality and impact. Microb Genomics 2022; 8: 1–10.

[24] Alcock BP, Huynh W, Chalil R, et al. CARD 2023: expanded curation, support for machine learning, and resistome prediction at the Comprehensive Antibiotic Resistance Database. Nucleic Acids Res 2023; 51: D690–D699.

[25] Rebelo AR, Bortolaia V, Leekitcharoenphon P, et al. One Day in Denmark: Comparison of Phenotypic and Genotypic Antimicrobial Susceptibility Testing in Bacterial Isolates From Clinical Settings. Front Microbiol 2022; 13: 1–12.

[26] Schwan CL, Lomonaco S, Bastos LM, et al. Genotypic and Phenotypic Characterization of Antimicrobial Resistance Profiles in Non-typhoidal Salmonella enterica Strains Isolated From Cambodian Informal Markets. Front Microbiol 2021; 12: 711472.

[27] Zhou Z, Alikhan NF, Mohamed K, et al. The EnteroBase user’s guide, with case studies on Salmonella transmissions, Yersinia pestis phylogeny, and Escherichia core genomic diversity. Genome Res 2020; 30: 138–152.

[28] Kille B, Nute MG, Huang V, et al. Parsnp 2.0: scalable core-genome alignment for massive microbial datasets. Bioinformatics 2024; 40: 0–4.

[29] Letunic I, Bork P. Interactive tree of life (iTOL) v5: An online tool for phylogenetic tree display and annotation. Nucleic Acids Res 2021; 49: W293–W296.

[30] Yang S, Xu H, Sun J, et al. Shifting trends and age distribution of ESKAPEEc resistance in bloodstream infection, Southwest China, 2012-2017. Antimicrob Resist Infect Control 2019; 8: 1–10.

[31] Lin JN, Lai CH, Yang CH, et al. Comparison of four automated microbiology systems with 16S rRNA gene sequencing for identification of Chryseobacterium and Elizabethkingia species. Sci Rep 2017; 7: 7–11.

[32] Phoenix □Automated Microbiology System User ‘ s Manua l. 003342.

[33] Id CNW, Id CVP, Akoko J, et al. Salmonella identified in pigs in Kenya and Malawi reveals the potential for zoonotic transmission in emerging pork markets. 2020; 1–16.

[34] Fàbrega A, Vila J. Salmonella enterica serovar Typhimurium skills to succeed in the host: Virulence and regulation. Clin Microbiol Rev 2013; 26: 308–341.

[35] Heredia N, García S. Animals as sources of food-borne pathogens: A review. Anim Nutr 2018; 4: 250–255.

[36] Dougan G, Baker S. Salmonella enterica serovar typhi and the pathogenesis of typhoid fever. Annu Rev Microbiol 2014; 68: 317–336.

[37] Kariuki S, Mbae C, Van Puyvelde S, et al. High relatedness of invasive multi-drug resistant non-typhoidal salmonella genotypes among patients and asymptomatic carriers in endemic informal settlements in kenya. PLoS Negl Trop Dis 2020; 14: 1–14.

[38] Kariuki S, Okoro C, Kiiru J, et al. Ceftriaxone-resistant Salmonella enterica serotype typhimurium sequence type 313 from Kenyan patients is associated with the blaCTX-M-15 gene on a novel IncHI2 plasmid. Antimicrob Agents Chemother 2015; 59: 3133–3139.

[39] Sora GH, Gachara G, Ichinose Y, et al. Antimicrobial Resistance Patterns of bacterial Septicaemia infecting infants in Mbita Subcounty, Western region of Kenya. J Nat Sci Res 2020; 10: 50–61.

[40] Zhan Z, Xu X, Gu Z, et al. Molecular epidemiology and antimicrobial resistance of invasive non-typhoidal salmonella in China, 2007–2016. Infect Drug Resist 2019; 12: 2885–2897.

[41] Kubicek-Sutherland JZ, Xie G, Shakya M, et al. Comparative genomic and phenotypic characterization of invasive non-typhoidal salmonella isolates from Siaya, Kenya. PLoS Negl Trop Dis 2021; 15: 1–26.

[42] Jang J, Hur HG, Sadowsky MJ, et al. Environmental Escherichia coli: ecology and public health implications—a review. J Appl Microbiol 2017; 123: 570–581.

[43] Kohli-Kochhar R, Omuse G, Revathi G. A ten-year review of neonatal bloodstream infections in a tertiary private hospital in Kenya. J Infect Dev Ctries 2011; 5: 799–803.

[44] Kibaba PW, Louis H, Kering KK, et al. Antimicrobal susceptibility pattern of methicillin resistant Staphylococcus aureus isolatetd from pediatric clinical samples at Webuye District Hospital. 2017; 74: 238–250.

[45] Lord J, Gikonyo A, Miwa A, et al. Antimicrobial resistance among Enterobacteriaceae, Staphylococcus aureus, and Pseudomonas spp. isolates from clinical specimens from a hospital in Nairobi, Kenya. PeerJ; 9. Epub ahead of print 2021. DOI: 10.7717/peerj.11958.

[46] Obanda BA, Gibbons CL, Fèvre EM, et al. Multi-Drug Resistant Staphylococcus aureus Carriage in Abattoir Workers in Busia, Kenya. Antibiotics 2022; 11: 1–14.

[47] Droz N, Hsia Y, Ellis S, et al. Bacterial pathogens and resistance causing community acquired paediatric bloodstream infections in low- And middle-income countries: A systematic review and meta-analysis. Antimicrob Resist Infect Control 2019; 8: 1–12.

[48] Orucho VO, Nyangau MK. Antibiotic resistance of Streptococcus pneumonia serotypes in Kisii, Kenya Cyrus Orucho Ochoi Kisii Teaching and referral hospital. 2020; 1–10.

